# Adolescents and young adults with cancer in Switzerland: a large single-centre analysis of key care pathways and outcomes by sex and age

**DOI:** 10.1101/2025.03.28.25324628

**Authors:** Pedersen Eva S L, Fischer Hanna J, Verschoor Daniëlle, Oramalu Ogo E, Ryser Christoph O, Kronig Marie-Noelle, Pabst Thomas, Berger Martin D, Segelov Eva, Özdemir Berna C

## Abstract

**Background:** Adolescents and young adults with cancer (AYA) represent a distinct patient group with unique challenges. Most studies reporting outcomes use registry data, leaving local care aspects underreported.

**Aim:** To understand demographic and clinical characteristics of AYA patients focusing on variations by cancer type, sex, and age, as well as fertility preservation, palliative care, clinical trial participation, and survival outcomes.

**Methods:** A retrospective single-centre study examined electronic medical records of AYA patients (age 15–39 years) treated for their initial diagnosis of cancer at University Hospital Bern (Inselspital) in the period 2015 -2021. Descriptive statistics summarized patient characteristics, and Kaplan-Meier survival curves and logistic regression were used for survival prediction.

**Results:** The AYA cancer cohort consisted of 395 patients (67% male) with a median age of 27. Testicular cancer and Hodgkin lymphoma were the commonest diagnoses (24% and 25% respectively), followed by non-Hodgkin lymphoma (11%), leukaemia (11%), and central nervous system tumours (11%). The median time from diagnosis to first treatment was 13 days. Clinical trial participation was 29% and 58% attended fertility consultations with higher rates among males (61%) than females (50%). Five-year survival for the entire cohort was 84%; lower in older patients and those with metastatic disease.

**Conclusion:** This study provides the first Swiss dataset on AYA cancer patients at a tertiary centre. While survival rates were high, disparities existed by diagnosis and metastatic status. Further research is needed to enhance survival in specific cancers and improve palliative and fertility care.

## Introduction

Adolescents and young adults (AYA), commonly defined as ages 15-39 years, with cancer is a distinct group of patients differing from children and older adults in terms of incidence, cancer biology, management needs and prognosis (1, 2). An estimated 120’000 AYA cancers were diagnosed in 2020 in Europe, corresponding to 5% of all new cases of cancer (3, 4). Globally among all people in this age group, cancer is the fourth leading cause of death (3, 4). Cancer incidence among AYA has increased since 1975 (5). Cancer in AYA patients affects more often females (60%) as compared to cancer in both children under 15 and adults aged 55 and older which predominantly affects males (6–9).

Cancer type in AYA varies by sex, where males have highest prevalence of thyroid (14%), testicular cancer (9%), and leukaemia (8%), compared to females with breast (29%), thyroid (21%) and cervix cancer (12%) (10–12). Survival is overall high, with cancer registry data from 29 countries among 700’000 AYA over the period 2010-2024 reporting five-year survival for all cancers of 84% (3). The best and worst outcomes were for Hodgkin lymphoma (HL) (95%) and acute lymphoblastic leukaemia (59%), respectively. Although an older series from the Surveillance, Epidemiology, and End Results (SEER) registry in the USA between 1975 and 2002 reported that survival rates among AYA improved less (average annual percent change (AAPC) 4.6%) over the time-period than children (AAPC 5.8%) (13), more recent data showed similar AAPC between 2000-2014 in survival for AYA (0.33%) and for children (0.36%) (14). There is conflicting data on whether the outcomes are worse for male or female patients (11, 15).

While extensive data exist for cancer among children and older adults, knowledge gaps remain for the AYA population. Most studies describing cancer in AYA are based on registry data which is excellent for describing epidemiological trends in prevalence, incidence, and prognosis, but information is lacking for local disease burden and care pathways, such as supportive care referrals for fertility preservation and palliative care. Few clinical studies exist, with most focusing on a single cancer type (16, 17) or including only limited data (18–21). This study presents a wholistic picture of AYA patients treated at the University Hospital Bern (Inselspital) over a recent six-year period to examine demographics, cancer characteristics, and patterns of care.

## Methods

### Study design and participants

This is a retrospective single-centre study using data extracted from electronic medical records from the Inselspital, University Hospital Bern, a tertiary referral centre covering the region of Bern, the capital of Switzerland, with around 1 million inhabitants. Individuals aged 15 to 39 at time of diagnosis treated for any cancer at adult oncological outpatient ward at the University Hospital Bern between January 1, 2015, and December 31, 2021, and who had signed the general informed consent form of the Inselspital (thereby allowing study entry) were included. The Ethics Committee of the Canton of Bern waived approval for this study (2022–00474). Patients with cancer diagnosed at younger age who presented with relapse as AYA were excluded, as were patients who started definitive cancer treatment at another centre.

### Data collection and definitions

Demographic and cancer characteristics were manually extracted from the hospital electronic medical system, IPDOS®. From this, the following parameters were calculated: age at diagnosis, body mass index (BMI (weight/height^2^), time between diagnosis (or if missing, referral date), and first consultation at Inselspital, five-year survival.

### Statistical analysis

Descriptive statistics were used to depict patient and cancer characteristics. Characteristics stratified by age, sex, and major diagnosis groups are shown where data sets include at least 10 patients, to preserve anonymity. Differences in age, BMI, first treatment, clinical trial participation, and five-year survival by major diagnosis groups were tested using K-sample equality of medians for non-normally distributed variables and chi^2^ for categorical variables. Kaplan-Meier plots were used to examine five-year survival. Predictors of survival were analysed by logistic regression analysis, with survival five years after follow-up used as the dependent variable and age at diagnosis, sex, and major diagnosis groups as independent variables. In a second model, we included a category of local versus metastatic disease at diagnosis for tumours where this categorisation is applicable, including testicular, Hodgkin lymphoma (HL), non-Hodgkin lymphoma (NHL), gastrointestinal (GI) cancers, and bone and soft tissue sarcoma. We present both models as the second model only includes a selection of the relevant diagnoses and results from this model cannot be generalized to all AYA in our sample. Analyses were computed using Stata 18 software, TX: StataCorp LLC for Windows.

## Results

Of 497 AYA with cancer who were treated at Inselspital Bern between January 2015 and December 2021 (**Supplementary table 1**), 395 received their definitive cancer treatment at Inselspital and were included in this analysis (67% males; median age 27 years; **Table 1)**. Testicular cancer and HL were the most common (24% and 25%, respectively), followed by NHL (11%), leukaemia (11%) and central nervous system (CNS) tumours (11%). Rare cancers (defined as diagnosed in less than five patients in our cohort) are presented in **Supplementary table 2.** Excluding testicular and female genital cancers, there was little difference in cancer origin between male and females.

**Table 1:** Characteristics of included patients (N=395)

|  | Male<br>N=264<br>n (%) | Female<br>N=131<br>n (%) | 15-19 y<br>N=69<br>n (%) | 20-29 y<br>N=205<br>n (%) | 30-39 y<br>N=121<br>n (%) | Total<br>N=395<br>n (%) |
| --- | --- | --- | --- | --- | --- | --- |
| **BMI at first treatment at Inselspital** |  |  |  |  |  |  |
| <18.5 | 10 (4) | 11 (8) | 9 (13) | 9 (4) | 3 (2) | 21 (5) |
| 18.5-24.9 | 163 (62) | 87 (66) | 46 (67) | 135 (66) | 69 (57) | 250 (63) |
| 25.0-29.9 | 67 (25) | 19 (15) | 10 (14) | 38 (19) | 38 (31) | 86 (22) |
| ≥30 | 22 (8) | 12 (9) | 3 (4) | 20 (10) | 11 (9) | 34 (9) |
| Unknown | 2 (1) | 2 (2) | 1 (1) | 3 (1) | 0 | 4 (1) |
| **Diagnosis** |  |  |  |  |  |  |
| Testicular cancer | 93 (35) | 0 | 4 (6) | 56 (27) | 33 (27) | 93 (24) |
| HL | 57 (22) | 42 (32) | 31 (45) | 52 (25) | 16 (13) | 99 (25) |
| NHL | 29 (11) | 16 (12) | 8 (12) | 17 (8) | 20 (17) | 45 (11) |
| Leukemia | 27 (10) | 15 (11) | 10 (14) | 16 (8) | 16 (13) | 42 (11) |
| CNS | 20 (8) | 23 (18) | 6 (9) | 24 (12) | 13 (11) | 43 (11) |
| Gastro-intestinal | 9 (3) | 5 (4) | 0 | 5 (2) | 9 (7) | 14 (4) |
| Bone and soft tissue | 15 (6) | 10 (8) | 5 (7) | 17 (8) | 3 (2) | 25 (6) |
| Breast and female genitals | 0 | 5 (4) | 0 | 1 (0) | 4 (3) | 5 (1) |
| Melanoma and other skin | 3 (1) | 0 | 0 | 3 (1) | 0 | 3 (1) |
| ENT | 4 (2) | 4 (3) | 2 (3) | 4 (2) | 2 (2) | 8 (2) |
| Urothelial | 1 (0) | 4 (3) | 2 (3) | 2 (1) | 1 (1) | 5 (1) |
| Other* | 6 (2) | 7 (5) | 1 (1) | 8 (4) | 4 (3) | 13 (3) |
| **Days between diagnosis and first treatment at Insel, median (IQR)**<br>(unknown for n=16) | 14 (7-29) | 11 (3-31) | 8 (1-18) | 14 (7-30) | 18 (7-44) | 13 (6-30) |
| **Included in clinical trials** | 71 (27) | 42 (32) | 21 (30) | 42 (20) | 50 (41) | 113 (29) |
| **Fertility consultation** | 162 (61) | 66 (50) | 40 (58) | 129 (63) | 59 (49) | 228 (58) |
| **Five-year survival (n=336#)** | 189 (85) | 94 (82) | 61 (95) | 159 (86) | 63 (72) | 283 (84) |
| **Palliative care consultation among patients who died (n=60)** | 16 (46) | 15 (60) | 1 (20) | 17 (59) | 13 (50) | 31 (52) |
| **Years between diagnosis and palliative care consultation (n=31), median (IQR)** | 1.3<br>(0.6-2.1) | 2.0<br>(1.1-3.1) | 11.3 | 1.7<br>(1.3-2.9) | 0.9<br>(0.5-2.1) | 1.6<br>(0.8-2.2) |

\*Other diagnosis described in detail in supplementary table 2. #Five-year survival calculated for 336 who were followed up for at least five years. +P-value calculated for test of equal medians and chi-squared for equal proportions. Abbreviations: HL; Hodgkin lymphoma, NHL; non-Hodgkin lymphoma, CNS; central nervous system, ENT; ear nose throat, BMI; body mass index, IQR; interquartile range, AYA; adolescents and young adults

Median time between diagnosis and first treatment was 13 days (interquartile range (IQR): 6-30). Clinical trial participation was recorded for 29% with higher participation rates for AYA aged 30-39 (41%) compared to younger AYA (15-19 year-olds 30% and 20-29 year-olds 20%). Further, trial participation was most common among patients with leukaemia, HL, and NHL and least common for testicular cancer and bone and soft tissue sarcoma (**Table 2**). Fertility consultation with a specialised service was document for 58% of patients (61% males and 50% females). The outcome of consultations is presented in **Figure 1**; more females than males rejected fertility preservation (26% and 8%, respectively).

**Table 2:**
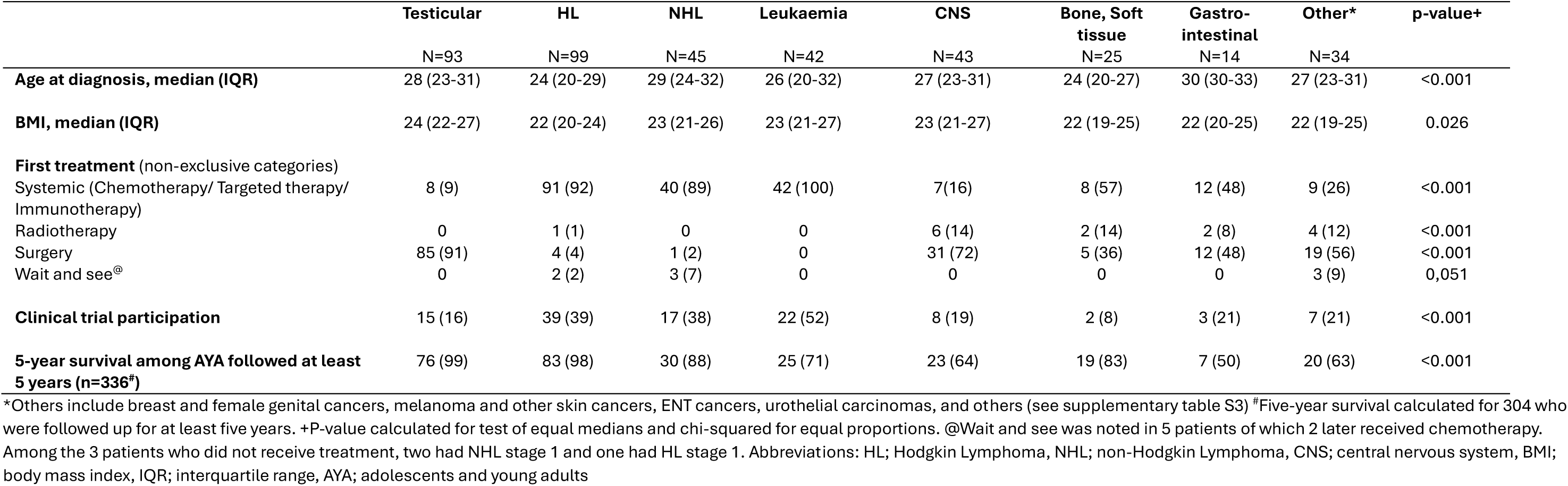
Diagnostic, treatment, and outcome characteristics by major diagnosis group (n=395)

**Figure 1:**
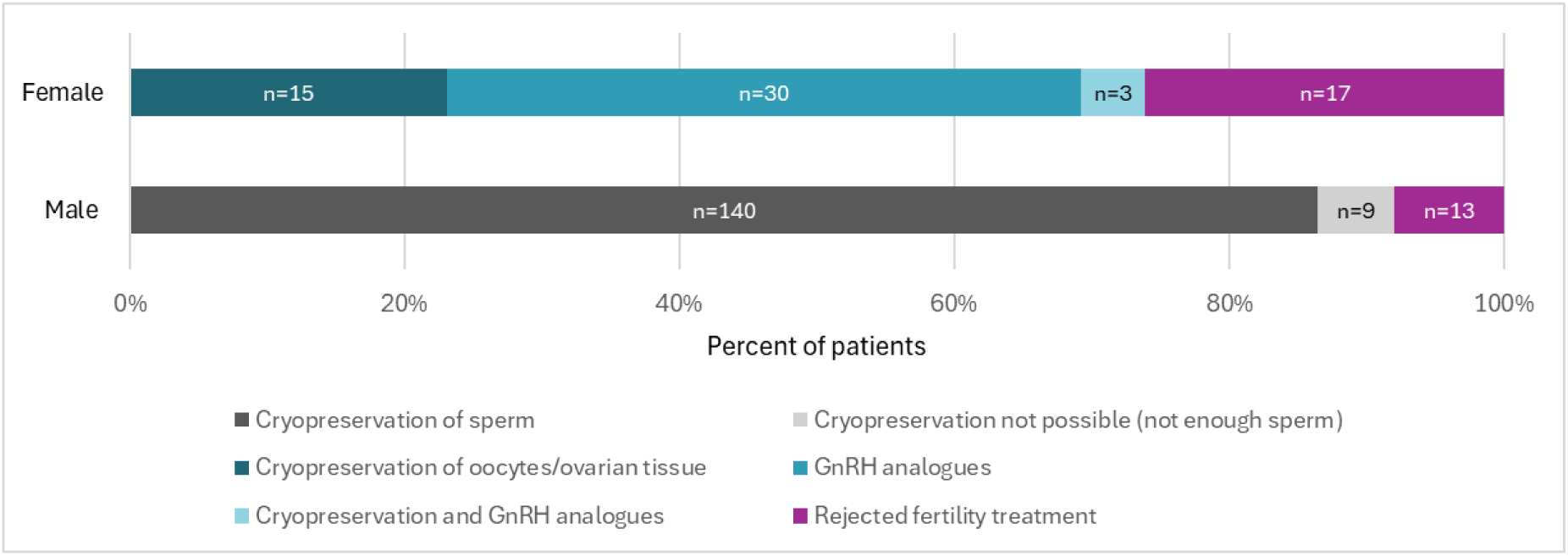
Fertility preservation among patients who had following fertility consultation (N=228)

Median age at diagnosis was lowest for HL (24 years) and bone and soft tissue sarcoma (24 years) and highest for the GI cancers (30 years) (**Table 2**). Stage of disease defined as localised or metastatic also differed between diagnoses, with most testicular cancers being localised while half of GI cancers were metastatic (**Figure 2**).

**Figure 2:**
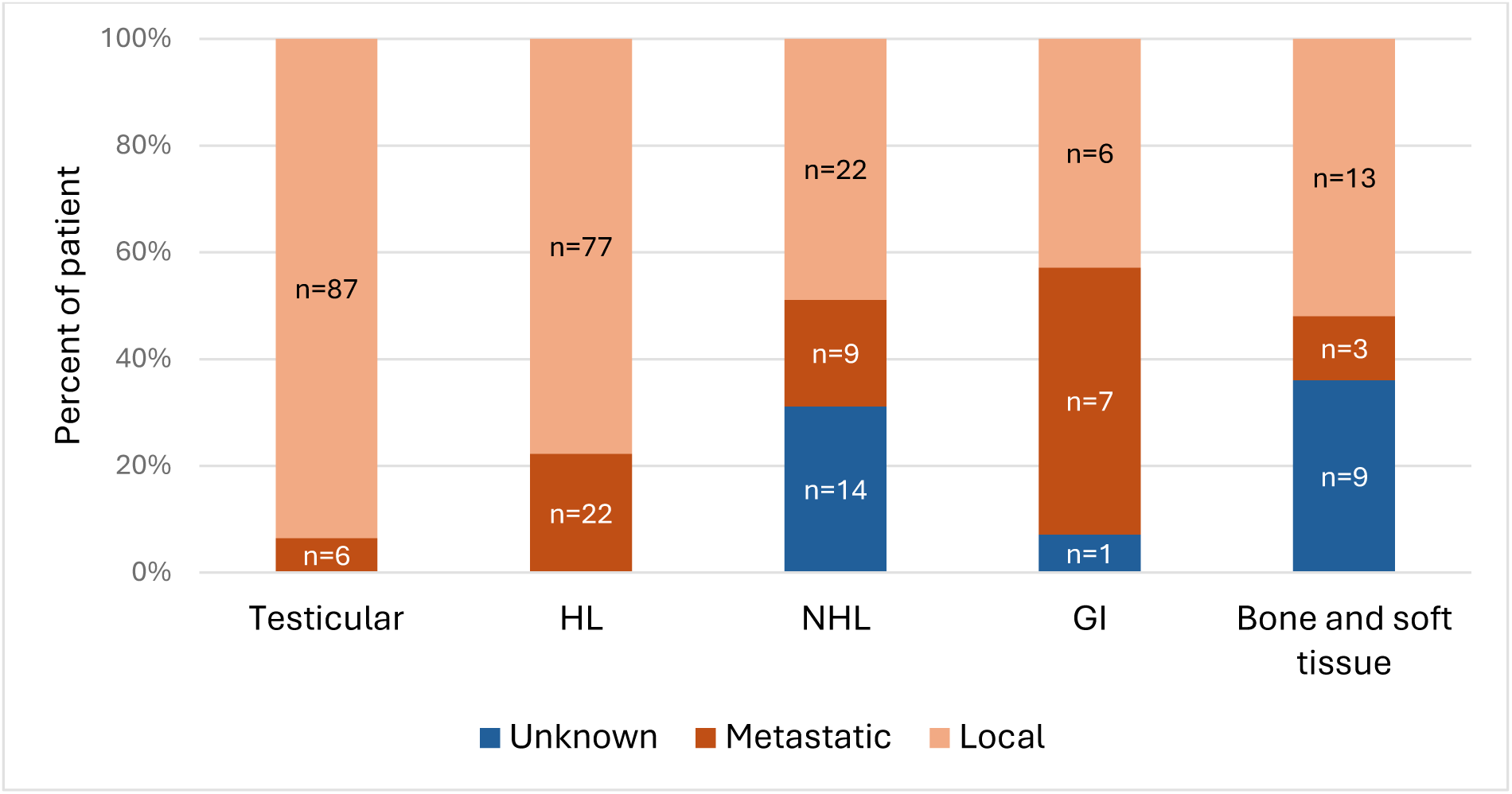
Local vs. metastatic disease for relevant diagnosis groups with at least 10 patients (N=276).

Five-year survival for the entire cohort was 84%, which decreased with older age. Highest survival was seen for testicular cancer (99%) and HL (98%) whilst poorest outcomes were for patients with GI cancer (50%) (**Table 2**, **Figure 3**). Lower odds of survival were associated with older age (Odds Ratio (OR): 0.88, 95% confidence interval (CI): 0.83-0.95) and diagnoses other than testicular cancer and HL (e.g. NHL with an OR of 0.08, 95% CI: 0.008-0.75), but not sex (**Figure 4A**). Using the model for the 238 patients whose tumours could be characterised as either localised or metastatic, diagnosis and metastatic disease (OR 0.23, 95%CI 0.08-0.66) were the only factors associated with lower survival (**Figure 4B**). Among patients, who died, 52% had a palliative care consultation with median years between diagnosis and palliative care consultation of 1.6 years (IQR: 0.8-2.2) (**Table 1**).

**Figure 3:**
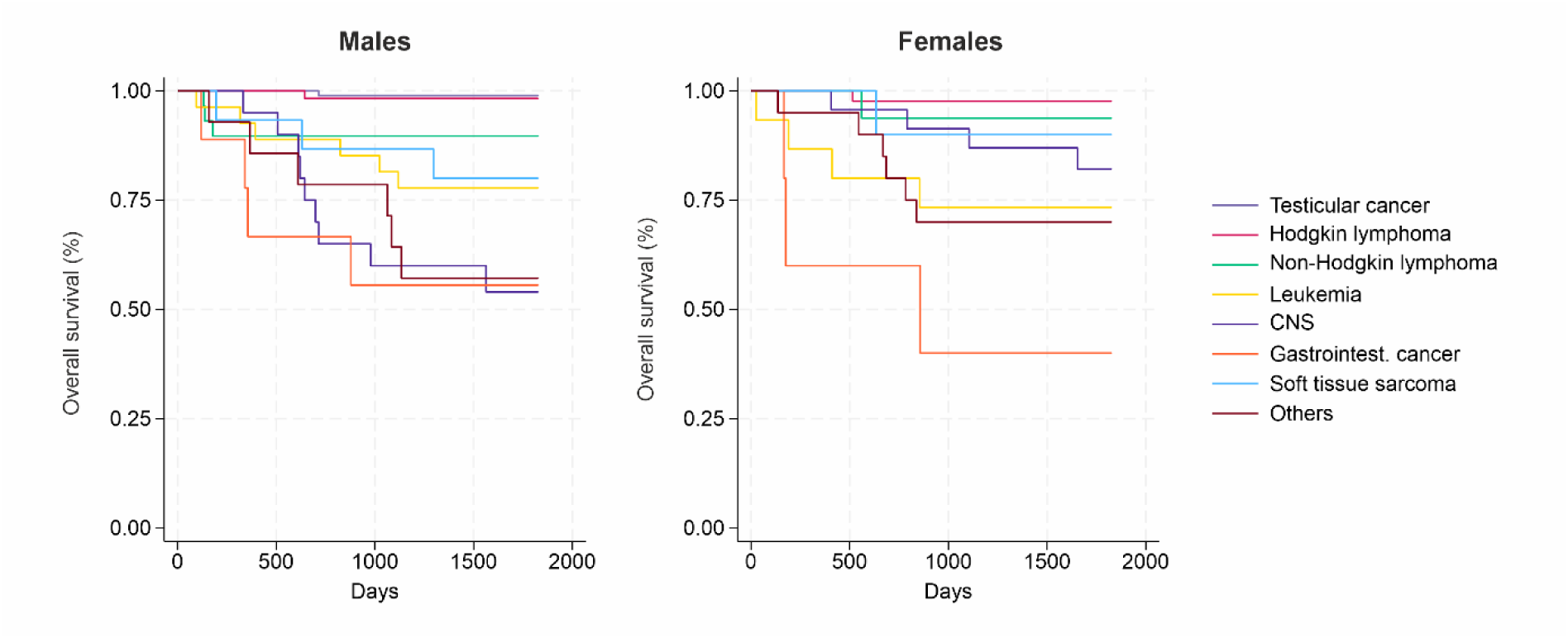
Kaplan Meier plots of 5-year survival for males and females

**Figure 4:**
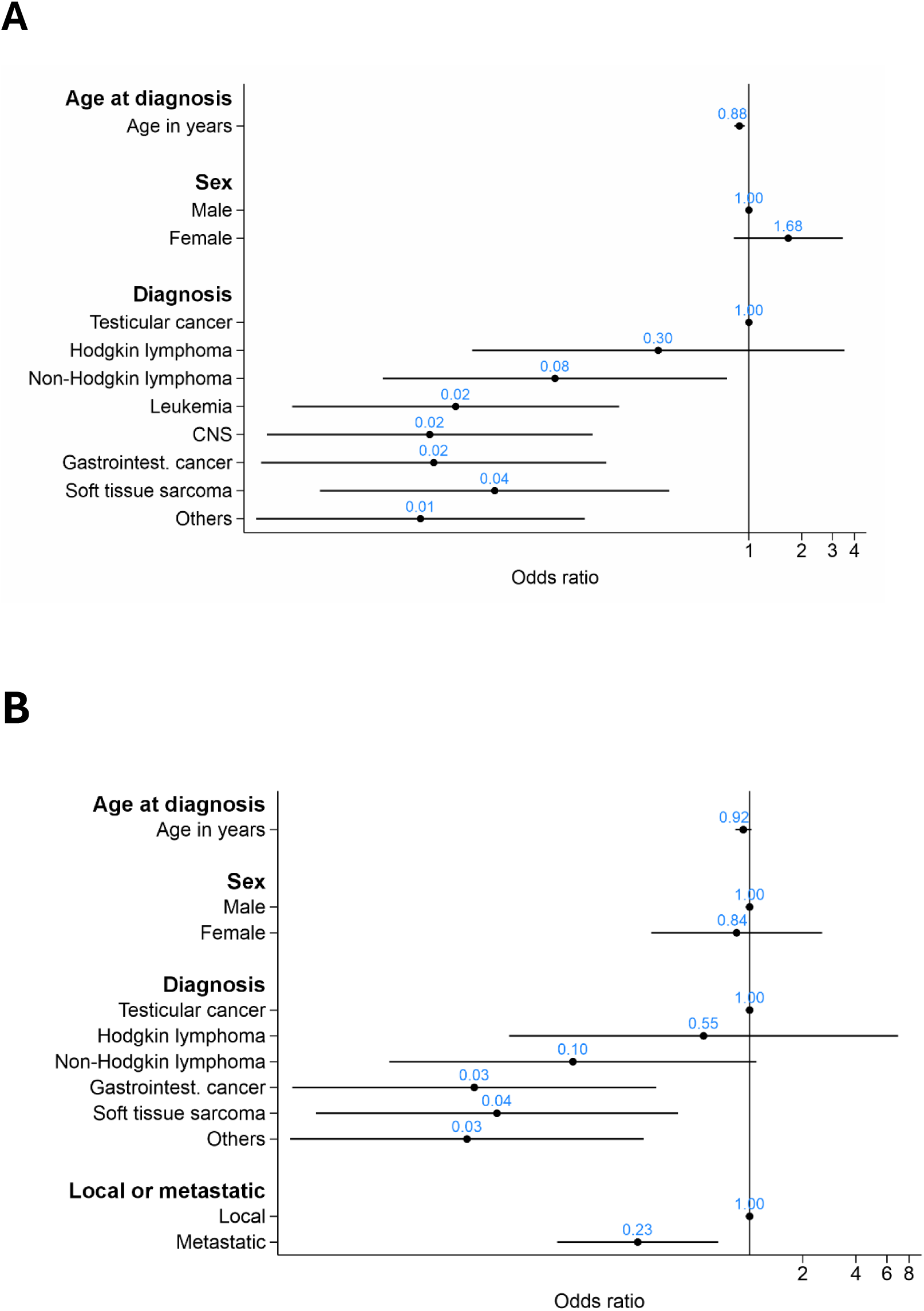
Results of logistic regression with survival at year 5 (yes/no) among people followed at least 5 years and A) including age at diagnosis, sex, and major diagnosis groups (N=336) B) including age at diagnosis, sex, major diagnosis groups, and local vs metastatic disease (N=238)

In 16 patients (4%), the cancer diagnosed and treated at Inselspital represented either second, distinct cancer, or a relapse of a previously known childhood cancer (**Supplementary Table 3**). The most common first diagnosis was leukaemia (n=6) and among 10 of 16, the second cancer was a recurrence. On average, the second cancer was diagnosed 3.4 years after the initial diagnosis. After a median follow-up of 7.7 year, 6 of these patients were deceased (37%), which is a higher proportion than among those with only one recorded tumour (14%).

## Discussion

This study presents the first Swiss data analysing the AYA population treated at one of six tertiary oncological referral centres. This comprehensive review of all patients treated over a 6-year period reflects an area of uptake of around one million people, representing around an eighth of the total contemporary Swiss population. While most published studies of AYA patients with cancer use registry data, this study shows the burden of disease and patterns of care from a clinical perspective. The few other existing clinical studies include either just one cancer type or present limited data, making it difficult to compare with our findings. Compared to registry series, there were some differences, particularly in distribution of cancer type and survival outcomes.

Just over half of the cancers in this series were HL and testicular cancer. By comparison, GLOBOCAN data from 2022 showed the highest prevalence of breast (19%) and thyroid cancer (18%) (11), while SEER data between 1975 and 1998 among AYA aged 20-29 years was more similar, with HL (12%), melanoma (12%), testicular (12%), and NHL (6%) (5). Among AYA 30-39 years old, breast cancer was most common (20%) followed by malignant melanoma (11%). An Australian study based on registry data from 2003-2015 also reported that 15% of all cancers in AYA were melanomas, but Australia has the highest rate of melanoma in the world with a age-standardized rate (ASR) per 100’000 of 35.1 (22), relating to its geography and weather (23). However, Switzerland also has a relatively high ASR of 20.3 (22) for melanomas, which is why it is unexpected to see so few cases in our sample. A possible explanation is that AYA diagnosed with melanoma are only referred to oncology centres in case of metastatic disease and that follow-up of localized melanoma is performed by dermatologists. The relatively low rate of breast cancer in our series is not explained by the overall breast cancer incidence in Switzerland, which is similar to other high income countries (24) but may be explained by the overall small numbers, and/or by referral patterns to private centres, although international guidelines recommend all AYA cancers be treated in a designated AYA centre.

Five-year survival was highest for testicular (99%) and HL (98%) and lowest for gastrointestinal cancer (50%) and CNS tumours (64%), which mirror the SEER AYA data from 2000-2014 showing five-year survival rate for HL of 94% and EUROCARE-6 data from 1999-2007 of 95% (3, 25). For testicular cancer, the EUROCARE-6 AYA data reported five-year survival rate of 97%, with 62% for CNS tumours (3).

With regards to the patterns of care, this study documented clinical trial participation at 29%, which is at the upper range of that reported in a recent review of 10 identified studies published between 2010 and 2018 of AYA clinical trial participation showing participation in 12% to 29% (26). To place this in context, trial participation in paediatric patients with cancer in the US is reported at 19% (27); by contrast, for adult cancer patients clinical trial participation rates ranged between 7.7% and 8.1% in different countries (28–30), with a similar rate of 7.8% clinical trial participation for adult patients receiving anti-cancer immunotherapies at our centre (31).

In our series, participation differed by diagnosis; more than half of patients with leukaemia participated in a trial, while for AYA with sarcoma and CNS tumours, respectively, this was only 8% and 19%. This difference is in line with existing evidence (26, 32) and most likely reflects complex factors that govern trial availability, including funding, clinical interest, and drug availability. Given that this is a retrospective analysis based on medical chart review, data relating to the number of patients who were offered a trial but declined to participate, or were ineligible, is not available. Studies examining clinical trial participation of AYA patients show that reasons for accepting or declining trial enrolment are multifactorial and related to patient, provider, and structural factors (33). For instance, most common reported reasons for nonparticipation include concerns about side effects of the treatment and concerns that the treatment had not been sufficiently tested (34).

Fertility is a major issue for patients with AYA cancers. The broad age range may include prepubertal individuals and those who have already fulfilled their fertility desires, but the majority are at a time in their life where options for fertility preservation are highly relevant. In this series, among those who had a fertility consultation, 75% of females and 92% of males had fertility preservation. This is substantially more than what was found in a study from another Swiss centre collecting data among patients treated for leukaemia between 2002 and 2012, in whom 1 of 44 (2.3%) female patients aged 17-45 years had fertility preservation and 33 of 101 (33%) male patients aged 14-60 years had fertility preservation (35). The lower rate of fertility preservation among females than males in our and previous studies maybe be because preservation is more complicated in females. It is unfortunate that most medical records do not require documentation of reasons fertility consultations were not undertaken, and we therefore do not know why some patients refused fertility preservation. However, one reason might be fear of treatment delay, but unlike previous decades, modern techniques not only mean higher rates of successful sperm or oocyte collection and subsequent usage for both men and women, but more rapid processes, meaning that these interventions usually no longer impact the timing of start of treatment (36). One further important factor is costs, as even for cancer patients, fertility preservation is not universally covered by health insurance in Switzerland (37, 38).

The referral to palliative care occurred in 8% of all patients, which is in line with recent data from a US tertiary centre including 4’674 AYA, where also 8% were referred to palliative care, mostly during inpatient treatment and more likely in specific cancer types with high symptom burden and/or poor survival (39). Unfortunately, due to low sample size, it was not possible to study differences in palliative care by diagnosis in our patient population. When considering only the patients who died in our cohort, 52% had palliative care, which is lower than the 90% palliative care rate among patients who died according to a population-based analysis of over 5’000 AYA patients in Ontario, Canada (40).

Strengths of our study include the broad inclusion of all AYA patients treated over a contemporary period at a large Swiss University hospital, compared to most studies in the literature which are based on registry data. This allows broader examination of patterns of care, as well as the ability to probe incomplete or unusual data through further examination of medical records. Study limitations include the cantonal nature of the Swiss healthcare system, which may affect the generalisability of the data. Complex AYA cancer patients are typically referred to specialised cancer centres like Inselspital, while the majority of AYA patients are still treated in peripheral oncology units. Furthermore, the sample size poses limitations for comparing characteristics between patients with less common tumours. While data on survival was carefully collected, there is potentially less rigorous data than a registry-based study. Finally, our sample might underrepresent those aged 15-17 years, as these might have been referred to the separate paediatric oncology centre.

This study underlies the critical move to identify AYA as a special group of patients with cancer, with particular physical and psychosocial care needs. The age range covers older patients with paediatric-type malignancies, younger patients with typically adult-age cancers, as well as the group of cancers that predominate in the AYA age range. In conclusion, our analysis shows that in this specialized centre, survival was high and in line with other series. Fertility preservation rates were higher than in other studies highlighting a possible improvement, but palliative care consultations were still scarce. Further research and reporting for this special group of patients is needed to ensure all receive the highest quality of comprehensive cancer care.

## Data Availability

Data in the present study are available upon reasonable request to the authors.

## Display items

**Supplementary table 1:** Characteristics of patients treated for cancer at Inselspital Bern between January 2015 and December 2021.

|  | **Male**<br>N=335 | **Female**<br>N=162 | **Total**<br>N=497 |
| --- | --- | --- | --- |
|  | n (%) | n (%) | n (%) |
| **Age at diagnosis** |  |  |  |
| 15-19 y | 46 (14) | 32 (16) | 78 (16) |
| 20-29 y | 183 (55) | 79 (49) | 262 (53) |
| 30-39 y | 106 (32) | 51 (31) | 157 (31) |
| **Diagnosis** |  |  |  |
| Testicular | 125 (37) | 0 | 125 (25) |
| HL | 62 (18) | 46 (28) | 108 (22) |
| NHL | 41 (12) | 23 (14) | 64 (13) |
| Leukemia | 34 (10) | 16 (10) | 50 (10) |
| CNS-Neoplasm | 21 (6) | 25 (15) | 46 (9) |
| Gastro-intestinal cancer | 12 (4) | 8 (5) | 20 (4) |
| Bone and soft tissue | 17 (5) | 13 (8) | 30 (6) |
| Breast and female genital | 0 | 14 (9) | 14 (3) |
| Melanoma and other skin | 9 (3) | 2 (1) | 11 (2) |
| ENT cancer | 4 (1) | 4 (2) | 8 (2) |
| Urothelial carcinoma | 2 (1) | 4 (3) | 6 (1) |
| Other* | 8 (2) | 7 (4) | 15 (3) |
| **First treatment** |  |  |  |
| Inselspital Bern | 264 (79) | 131 (81) | 395 (79) |
| Other Swiss hospital | 55 (16) | 23 (14) | 78 (16) |
| Hospital in another country | 10 (3) | 3 (2) | 13 (3) |
| Unknown | 6 (2) | 5 (3) | 11 (2) |

\*Other diagnoses displayed in detail in supplementary table 2. Abbreviations: . Abbreviations: HL; Hodgkin lymphoma, NHL; non-Hodgkin lymphoma, CNS; central nervous system, ENT; ear nose throat, BMI: body mass index

**Supplementary table 2:** Exact diagnosis among patients with gastro-intestinal, breast and other female genital tract, and other cancers

|  | Male | female |
| --- | --- | --- |
| **Gastro-intestinal** | **9** | **5** |
| Colon carcinoma | 3 | 2 |
| Gastroesophageal cancer | 2 | 1 |
| Hepatocellular and cholangiocarcinoma | 1 | 0 |
| Pancreatic carcinoma | 1 | 0 |
| Rectal carcinoma | 2 | 2 |
| **Breast and female genital** |  | **5** |
| Breast cancer ER and PR positive HER2 negative |  | 1 |
| Breast cancer triple negative |  | 4 |
| **Other** | **6** | **7** |
| **Carcinoid** |  |  |
| Neuroendocrine tumor of the cervix |  | 1 |
| Atypical carcinoid of the lung with pulmonary, hepatic, and osseous metastases | 1 |  |
| **Blood vessel tumor (rare vascular malignancies)** |  |  |
| Metastatic malignant endothelial neoplasia | 1 |  |
| Hepatic und pulmonary hemangioendothelioma |  | 1 |
| Metastatic angiosarcoma of the right atrium | 1 |  |
| **Langerhans cell histiocytosis** |  |  |
| Multisystem Langerhans cell histiocytosis | 1 |  |
| Monosystem, unifocal Langerhans cell histiocytosis |  | 1 |
| Langerhans cell histiocytosis, eosinophilic granuloma | 1 |  |
| Multifocal Langerhans cell histiocytosis |  | 1 |
| **Other** |  |  |
| Adenocarcinoma of the lung with lymphatic and osseous metastases |  | 1 |
| Myeloproliferative hypereosinophilic syndrome |  | 1 |
| Neuroendocrine tumor of the thymus with lymphatic, pulmonary, and hepatic metastases |  | 1 |
| Non-Langerhans cell histiocytosis | 1 |  |

**Supplementary table 3:** Characteristics among patients with a second cancer treated at Inselspital Bern

| Patient number | Diagnosis 1 | Diagnosis 2 | Years between diagnoses | Recurrence or second primary | Alive |
| --- | --- | --- | --- | --- | --- |
| 1 | Squamous cell carcinoma of the skin | Lymphatic metastasis | 3.0 | Recurrent | No |
| 2 | Seminoma on the left side | HL | 5.0 | Second primary | Yes |
| 3 | B-ALL | CNS recurrence in B-ALL | 2.6 | Recurrent | Yes |
| 4 | Myelodysplastic syndrome (RAEB-1) | Secondary AML | 2.8 | Recurrent | No |
| 5 | Non-seminomatous testicular cancer on the right | Non-seminomatous testicular cancer on the left | 8.9 | Second primary | Yes |
| 6 | B-ALL | Recurrence in the right testicle | 11.1 | Recurrent | Yes |
| 7 | Metastatic pleomorphic rhabdomyosarcoma from a germ cell tumor | Mixed seminomatous/non-seminomatous testicular cancer | 3.5 | Second primary | No |
| 8 | B-ALL | Recurrence in the right testicle | 2.9 | Recurrent | Yes |
| 9 | Squamous cell carcinoma of the left ear helix | Multiple non-melanoma skin cancers, with history of Xeroderma Pigmentosum | 2.5 | Second primary | No |
| 10 | Nodular sclerosing HL | HL recurrence | 2.7 | Recurrent | Yes |
| 11 | B-ALL | B-ALL recurrence | 2.0 | Recurrent | Yes |
| 12 | Nodular sclerosing HL | Pleomorphic liposarcoma of the right scalenus medius muscle | 2.8 | Second primary | Yes |
| 13 | AML | Early extramedullary leukemia recurrence as myelosarcoma | 0.3 | Recurrent | No |
| 14 | Epithelioid hemangioendothelioma with hepatic and pulmonary metastasis | Non-ulcerated malignant melanoma of the superficial spreading melanoma type | 1.7 | Second primary | Yes |
| 15 | AML | AML recurrence | 0.9 | Recurrent | No |
| 16 | Follicular non-Hodgkin lymphoma | NHL recurrence | 9.8 | Recurrent | Yes |

Abbreviations: HL; Hodgkin Lymphoma, B-ALL; B Cell Acute Lymphoblastic Leukemia, CNS; central nervous system, AML; Acute myeloid leukemia, NHL; Non-Hodgkin Lymphoma.

